# Identifying individuals at risk for surgical supravalvar aortic stenosis by polygenic risk score with graded phenotyping

**DOI:** 10.1101/2024.09.17.24313555

**Authors:** D. Liu, C.B. Mervis, M.D. Levin, E. Biamino, M.F. Bedeschi, M.C. Digilio, G.M. Squeo, R. Villa, S. Osgood, J.A. Freeman, N. Raja, G. Merla, A.E. Roberts, C.A. Morris, L.R. Osborne, B.A. Kozel

## Abstract

In a previous pathway-based, extreme phenotype study, we identified 1064 variants associated with supravalvar aortic stenosis (SVAS) severity in people with Williams syndrome (WS) and either no SVAS or surgical SVAS. Here, we use those variants to develop and test polygenic risk scores (PRS). We used the clumping and thresholding (CT) approach on the full 1064 variants and a 427-variant subset that was part of 13 biologically relevant pathways identified in the previous study. We also used a lasso approach on the full set. We were able to achieve an area under the curve (AUC) of >0.99 for the two CT PRS methods, using only 622 and 320 variants respectively when 2/3 of the initial 217 participants data were used for training and 1/3 for testing. The lasso performed less well. We then evaluated the performance of those PRS variant sets on an additional group of 138 patients with WS with intermediate severity SVAS and found a misclassification rate of <10% between the surgical and intermediate groups, suggesting potential for clinical utility of the score.

---

Supravalvar aortic stenosis (SVAS) is a characteristic feature of Williams-Beuren syndrome (WBS). It can be “discrete” (characteristically seen as an hourglass narrowing in the supravalvar region of the aorta) or “diffuse” (sometimes referred to as long segment hypoplasia). Roughly 20% of people with WBS have discrete SVAS requiring surgical intervention in early childhood, while ∼35% have no significant discrete stenosis (hereafter, “no SVAS”), and ∼45% have mild or moderate discrete aortic narrowing. Until now a clinically usable genetic scoring system, such as polygenic risk score (PRS), for early identification of children with WBS at risk of surgical SVAS has not been available. In our recent modifier study^1^ evaluating 217 individuals with WBS (87 with surgical discrete SVAS and 130 with no SVAS, two extreme groups), we identified 427 common non-synonymous variants (hereafter variants) in 360 genes that were part of 13 key pathways associated with extreme SVAS outcomes from a total of 1064 originally selected variants with CADD>10 and a >5% difference in cohort-based allele frequency (Figure 1A). These pathways include innate/adaptive immune and matrisome. One challenge in genomic/genetic studies on both rare and common conditions is to build highly predictive PRS models with small variant sets for both clinical applications and mechanistic insight.^2^ In this letter, we ask whether PRSs calculated from either the 427 key pathway variants and/or the full set of 1064 variants are sensitive for classification of surgical SVAS vs. no SVAS phenotype by area under the curve (AUC) of receiver operating characteristics. We then evaluate the distribution of predicted PRSs for 138 additional individuals with mild/moderate SVAS using each of the two sets of variants. Finally, we evaluate whether pathway-based variant selection performs better in terms of sparsity of variants than Lasso-based methods.

**Figure 1.**
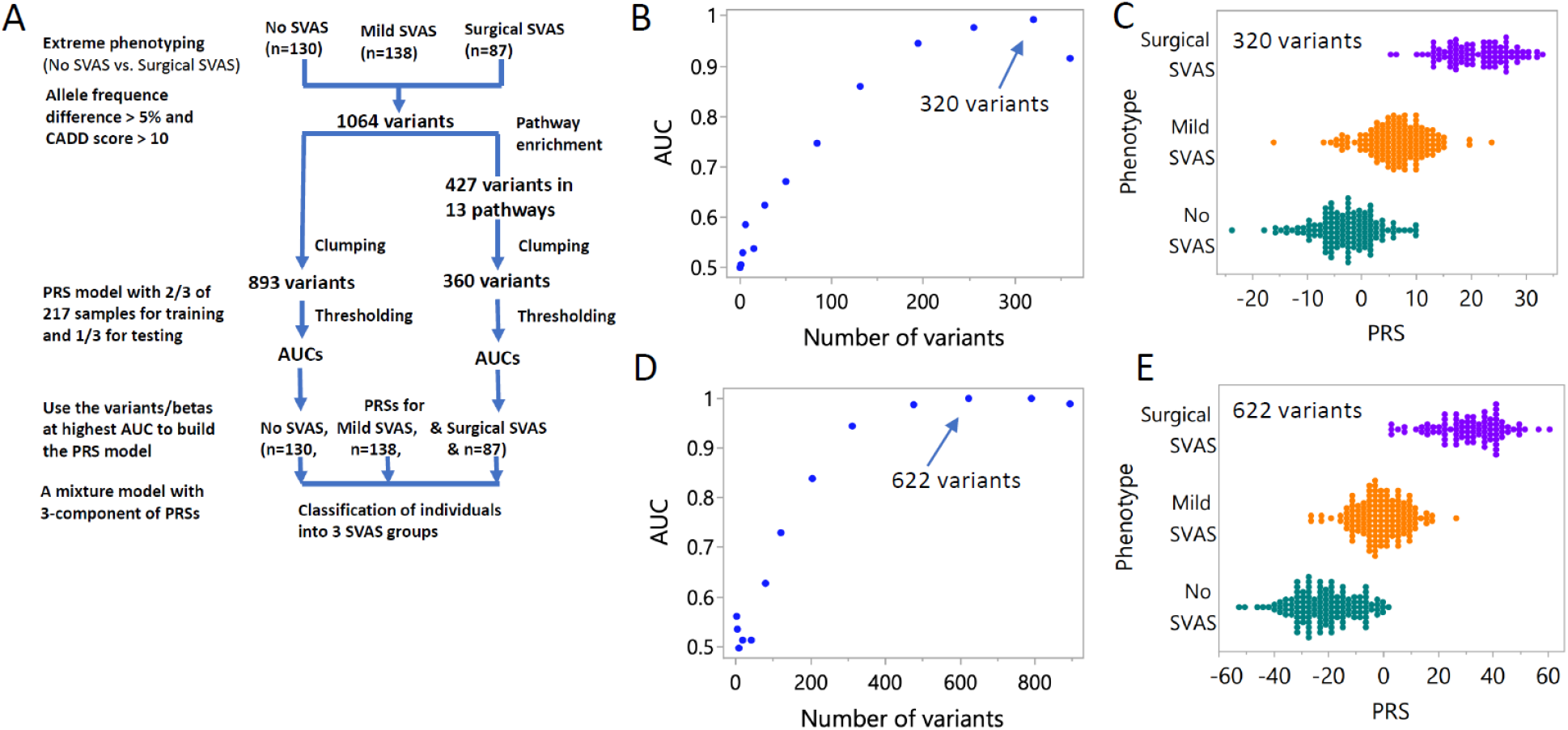
Flow diagram of the PRS calculations, the classification accuracy by AUC of surgical SVAS vs no SVAS, and predicted PRSs for the mild/moderate SVAS (n=138). **A**, Flow diagram of the PRS calculations for the “full” and “pathway enrichment” approaches. The median (25^th^, 75^th^ percentile) age (years) of last phenotyping in the no SVAS, mild/moderate SVAS and surgical SVAS subgroups of 355 individuals are 14.0 (8.1, 26.3), 9.6 (5.7, 16.0) and 10.5 (4.0, 19.0), respectively. **B**, Plot of AUC vs number of variants selected by -log10 p-value between 0 and 3 by 0.25 for the set of 360 variants in 13 previously defined key pathways^1^. **C**, Distribution of PRSs (not scaled) of 87 surgical SVAS (purple) and 130 no SVAS (dark green) cases calculated from 320 of the 360 variants, corresponding to AUC = 0.992. The PRSs of the 138 mild/moderate SVAS (orange) cases were computed with the same betas and 320 variants (arrow). Median (25^th^, 75^th^ percentile) PRSs of the three groups: no SVAS, mild/moderate SVAS and surgical SVAS in panel **C** are -2.6 (−6.5, 0.7), 6.2 (2.8, 9.5) and 20.4 (16.2, 25.4), respectively. **D**, Plot of AUC vs number of variants selected by -log10 p-value between 0 and 3 by 0.25 for the full set of 893 variants. **E**, Distribution of PRSs (not scaled) in 87 surgical SVAS (purple) and in 130 no SVAS (dark green) cases, calculated from 622 of the 893 variants, corresponding to AUC = 1.0 (arrow). The PRSs of the 138 mild/moderate SVAS (in orange) cases were computed with the same betas and 622 variants. Median (25^th^, 75^th^ percentile) PRSs of the no SVAS, mild SVAS and surgical SVAS groups in panel **E** are -22.8 (−30.9, -14.3), -1.0 (−6.8, 4.7) and 31.8 (22.3, 39.6), respectively.

See Liu et al^1^ for complete study demographics and participant consent information. Of the 184 individuals with mild/moderate SVAS originally reported, 46 were < 3 years of age at last phenotyping. As in the previous analysis, children < 3 years old who had not yet had surgery were excluded from analysis due to the potential for SVAS to progress in that window. About 92.1% of the participants exhibited Non-Finnish European ancestry, 1.4% were Asian, and 6.5% were of mixed ancestry. We took the clumping and thresholding (CT) approach using the bigsnpr package^3^ to estimate the betas via univariate logistic regression of the extreme outcomes: surgical SVAS and no SVAS for each of the 360 enrichment variants (after clumping out 67 variants), computed the PRSs for a randomly selected 2/3 of the 217 individuals (training data), and then tested the classification accuracy of the binary outcomes by AUC using the remaining 1/3 of the group (testing data) via 13 thresholds (number of variants) (Figure 1B). The distribution of the PRSs calculated using these 320 variants, corresponding to the highest AUC (0.992), is shown in Figure 1C (purple and green). We repeated the steps above for the full dataset of 1064 variants (893 after clumping out 171), and plotted AUCs vs the number of variants at the same 13 thresholds (Figure 1D). With inclusion of 622 of the 893 variants, the corresponding best AUC is 1.0. The distribution of the PRSs calculated with these 622 variants is shown in Figure 1E. The classification performance by AUC using the PRS scores calculated from the pathway-based list is almost identical to the full set for identifying the individuals at risk of surgical SVAS (Figures 1B and D). Importantly, the 320 pathway-based variants are more biologically relevant than the full set to the disease process.

We then estimated the PRSs using the betas, corresponding to AUC = 0.992 and AUC = 1.0 with 320/360 variants and 622/893 variants, respectively for the 138 individuals with mild/moderate SVAS, whose data were not part of the extreme phenotype model in our previous modifier study. The implicit assumption made here is that the effect of each variant (beta) in the mild/moderate SVAS group and surgical SVAS group relative to the no SVAS group is the same, which is in parallel with the proportional hazard assumption in the Cox proportional hazard model. The distributions of the PRSs calculated with the set of 320/360 “pathway” variants and the “full” set of 622/893 variants are shown in panels C and E in orange. The finding that the distribution of PRSs fell into three groups establishes mild/moderate SVAS as a phenotypic entity that lies on a genetic spectrum intermediate between the no SVAS and surgical SVAS groups. To estimate the classification error by PRS between the mild/moderate SVAS and surgical SVAS groups, we fit the re-sampled PRSs with replacement, calculated with 320 “pathway” and 622 “full” model variants for the three phenotype groups with a Gaussian 3-component mixture model 1000 times. The median classification errors (25^th^, 75^th^) for mild/moderate SVAS *vs*. surgical SVAS are similar at 0.095 (0.080, 0.11) and 0.069 (0.057, 0.081), respectively. The median classification errors (25^th^, 75^th^) for no SVAS *vs*. mild/moderate SVAS are 0.19 (0.17, 0.21) and 0.16 (0.14, 0.17), respectively.

Finally, we also examined whether lasso-based methods, such as lassosum2 in the bigsnpr package and the glmnet R package could achieve AUC > 0.9 with a sparse set of variants (e.g., 320 variants) from the “full” data pool. Ultimately, lasso-based methods were not able to reduce the full 893 variant set (after clumping) to a subset of similar size to the pathway-based approach (∼350) without compromising high AUC value (The ∼825 lasso-selected variants showed an AUC of 0.90 with penalty alpha = 0.05 in Lasso of the glmnet package.), highlighting the benefit of “pathway”-based variant model.

In those with WBS, surgical SVAS is normally diagnosed early in childhood, implying a strong genetic contribution. It is not surprising to observe a high AUC in that setting. In contrast, the published AUCs for more common complex cardiometabolic conditions are in the range of 0.6 and 0.84^4^, requiring 1000s to 100,000s of variants for the calculation of the PRS. While most PRS studies focus on cases and controls, ours is the first to consider a 3^rd^ intermediately affected group. These mildly affected people score between the extreme phenotype groups using as few as 320 variants, suggesting that mild/moderate disease is a both phenotypically and genetically separable subtype. The larger overlap on the no SVAS-mild/moderate side likely is partially due to imprecision in measurement, as those with milder disease have fewer and less invasive/definitive imaging studies. Additional studies incorporating differences between the mild/moderate and the two extreme groups are needed to identify specific pathways/variants that more precisely differentiate mild/moderate SVAS from surgical SVAS or no SVAS.

In summary, our extreme phenotyping method provided useful variants for construction of a highly effective PRS using data from relatively small numbers of patients for input. The “pathway” and “full” models performed similarly but the pathway method required fewer variants for its high AUC and has the added benefit of incorporating variants that are part of biologically relevant pathways. A similar two-tiered (extreme phenotype-based PRS followed by application to the larger intermediate dataset) approach may be employed in a variety of studies to limit sequencing cost. The insights of our approach can then be used to build more sensitive classifiers with PRSs and clinical risk factors for complex diseases^5^.

## Data Availability

All data produced in the present study are available upon reasonable request to the authors

## Acknowledgments

We would like to thank the individuals with WBS and their families for their participation in our study. Without their participation, this study would not have been possible. The NIH effort was supported by the NHLBI Division of Intramural Research (BAK). CAM was supported by grants from the Williams Syndrome Association (WSA) and LRO received funding from the Canadian Institutes for Health Research (MOP77720). CBM received support from the National Institute of Neurological Disorders and Stroke (R01 NS35102) and the WSA (WSA 0104 and WSA 0111). CAM and LRO were also partially supported by subcontracts from R01 NS35102. The Genomic Disorder Biobank of the Telethon Network of Genetic Biobanks was supported by Telethon Italy grant GTB12001G to GM. We thank Barbara R. Pober for contributing DNA samples to our study on WBS.

